# Methods for cost-efficient, whole genome sequencing surveillance for enhanced detection of outbreaks in a hospital setting

**DOI:** 10.1101/2024.02.16.24302955

**Authors:** Kady D. Waggle, Marissa Pacey Griffith, Alecia B. Rokes, Vatsala Rangachar Srinivasa, Erin M. Nawrocki, Deena Ereifej, Rose Patrick, Hunter Coyle, Shurmin Chaudhary, Nathan J. Raabe, Kathleen Shutt, Alexander J. Sundermann, Vaughn S. Cooper, Lee H. Harrison, Lora Lee Pless

**Affiliations:** Microbial Genomic Epidemiology Laboratory, Center for Genomic Epidemiology, University of Pittsburgh, 3507 Victoria Street, Pittsburgh, Pennsylvania, USA; Division of Infectious Diseases, University of Pittsburgh School of Medicine, 3550 Terrace Street, 818 Scaife Hall, Pittsburgh, Pennsylvania, USA; Department of Epidemiology, School of Public Health, University of Pittsburgh, 130 De Soto Street, Pittsburgh, Pennsylvania, USA; Department of Infectious Diseases and Microbiology, School of Public Health, University of Pittsburgh, 130 De Soto Street, Pittsburgh, Pennsylvania, USA; Department of Microbiology and Molecular Genetics, University of Pittsburgh School of Medicine, Pittsburgh, PA, USA; Center for Evolutionary Biology and Medicine, University of Pittsburgh School of Medicine, Pittsburgh, PA, USA

**Author notes:** Corresponding author and email address Lora Lee Pless University of Pittsburgh, Starzl Biomedical Science Tower 200 Lothrop St, Pittsburgh, PA 15261.

**Keywords:** Healthcare-Associated Transmission, Bacteria, Antimicrobial Resistance, Whole Genome Sequencing, Laboratory Methods, Cost Analysis

## Abstract

**Introduction:** Outbreaks of healthcare-associated infections (HAI) result in substantial patient morbidity and mortality; mitigation efforts by infection prevention teams have the potential to curb outbreaks and prevent transmission to additional patients. The incorporation of whole genome sequencing (WGS) surveillance of suspected high-risk pathogens often identifies outbreaks that are not detected by traditional infection prevention methods and provides evidence for transmission. Our approach to real-time WGS surveillance, the Enhanced Detection System for Healthcare-Associated Transmission (EDS-HAT), has 1) identified serious outbreaks that were otherwise undetected and 2) shown the potential to be cost saving.

**Methods:** We describe our cost-efficient methods to perform WGS surveillance and data analysis of pathogens for institutions that are interested to expand infection prevention surveillance. We provide an overview of the weekly workflow of EDS-HAT during two distinct phases over three years.

**Results:** In an average week at our tertiary healthcare system, we sequenced 60 samples at a cost of less than $100 each during Phase 1, and 80 samples for less than $70 each in Phase 2, inclusive of laboratory reagents and staff salaries. The average turnaround time, from sample collection to reporting data to infection prevention, was ten days.

**Conclusions:** Performing EDS-HAT in real-time can be both feasible and time-efficient. Providing such timely information to aid in outbreak detection could identify transmission events sooner and thus could increase patient safety.

**Impact statement:** Whole genome sequencing (WGS) surveillance to confirm or refute suspected outbreaks of potential healthcare-associated infections (HAI) is a highly effective approach for outbreak detection. Since November 2021, we have conducted WGS surveillance in real-time through a program called the Enhanced Detection System for Hospital-Associated Transmission (EDS-HAT), to assist our hospital infection prevention and control (IP&C) team to identify and stop outbreaks. Our laboratory has successfully implemented real-time WGS surveillance of multiple pathogens in the hospital setting continuously for over four years. Our weekly workflow included identifying HAI pathogens and performing WGS, followed by bioinformatic analyses that included species confirmation, determination of sequence type, and genetic relatedness comparisons. Based on this information, transmission clusters were identified, and the electronic health record was reviewed to determine probable transmission routes. Finally, IP&C implemented appropriate interventions to mitigate the spread of infection. The focus of this manuscript is to provide the details of our laboratory and analytical methods, along with the cost associated with laboratory materials and staff salary, for successful implementation of real-time WGS surveillance.

**Data Summary:** The whole genome sequencing data generated in this study are deposited in the United States National Institutes of Health, National Library of Medicine (https://www.ncbi.nlm.nih.gov/bioproject), and are publicly available under BioProject accession PRJNA475751. All supporting data and protocols are provided within the article or through supplementary data files.

## Introduction

Healthcare-associated infections (HAIs) are associated with substantial morbidity and mortality. HAIs also impose a significant economic burden on healthcare systems, costing hospitals an estimated $9.6 billion USD/ year(1, 2). Whole genome sequencing (WGS) for HAI organisms can provide insight on the transmission dynamics in hospital settings(3–5). Historically, determining the degree of genomic relatedness between organisms was accomplished using pulsed field gel electrophoresis (PFGE)(6). Given its many advantages and recent decline in cost, WGS has emerged as the leading method for determining genetic relatedness between clinical isolates(7, 8). Few hospital systems perform prospective WGS routinely or for an extended time; thus, reactive sequencing is often used to confirm or refute the presence of a suspected outbreak for most hospitals. This approach can fail to detect important outbreaks for a variety of reasons, including outbreaks caused by common organisms, those not clustering on a single nursing unit, those consisting of a small number of patients, or those caused by an unsuspected and/or complex transmission route(9–11). Furthermore, WGS can be used to infer phylogenetic relationship among organisms, detect the presence of antimicrobial resistance (AMR) genes and mobile genetic elements, and identify rare and/or novel genetic variants(10–14). The Microbial Genomic Epidemiology Laboratory (MiGEL) at the University of Pittsburgh developed the Enhanced Detection System for Healthcare-Associated Transmission (EDS-HAT) to identify outbreaks of HAIs in real time using WGS surveillance methods in partnership with the UPMC IP&C team and the UPMC Clinical Laboratories. EDS-HAT is currently in operation at our institution and has been for over four years(10, 11, 14–17).

Presumed barriers for most hospital systems for implementing WGS surveillance include cost, lack of technical expertise, and inadequate infrastructure. In this paper, we describe our laboratory methods, bioinformatics workflow, and provide a cost estimate of WGS surveillance, with the goal of providing guidance to hospitals that seek to implement WGS surveillance.

## Materials and Methods

### Study setting

MiGEL is a non-Clinical Laboratory Improvement Amendments (CLIA) certified research laboratory located on the University of Pittsburgh main campus, in Pittsburgh PA, USA. EDS-HAT was developed and is currently implemented in real-time at MiGEL in coordination with the University of Pittsburgh, UPMC, the UPMC Clinical Laboratory Building (CLB) team, the UPMC IP&C team, and Carnegie Melon University (CMU). UPMC Presbyterian is an adult tertiary acute care hospital with 758 total beds, 134 critical care beds, and over 400 annual solid organ transplants. UPMC Presbyterian Hospital, the primary teaching hospital of the system, is located adjacent to the University of Pittsburgh’s main campus. The University of Pittsburgh Institutional Review Board provided ethics approval for EDS-HAT (Protocol: STUDY21040126).

### Clinical specimen collection

#### Isolate inclusion criteria

A list of select, high-concern bacterial pathogens from UPMC Presbyterian Hospital was generated twice weekly using Theradoc (5.4.0.HF1.102, Pittsburgh, PA; Fig 1A). Pathogens of interest included: extended-spectrum beta-lactamase-producing (ESBL) *Escherichia coli,* ESBL *Enterobacter* species, *Acinetobacter* species, *Pseudomonas* species, *Klebsiella* species, *Stenotrophomonas* species, *Serratia* species, *Burkholderia* species, *Providencia* species, *Proteus* species, *Citrobacter* species, vancomycin-resistant *Enterococcus* (VRE), methicillin-resistant *Staphylococcus aureus* (MRSA), and *Clostridioides difficile*. EDS-HAT isolate inclusion criteria required patients to have been hospitalized for ≥3 days and/or to have had a previous hospital exposure within the 30-days prior to culture(10). In this study, we describe the samples and methods utilized to perform real-time EDS-HAT from March 2022-March 2025. We have described two phases, which were defined by a transition from the use of our initial sequencing instrumentation (MiSeq and NextSeq550) to more recent, higher throughput technologies (NextSeq1000 and NovaSeq X Plus).

**Fig 1.**
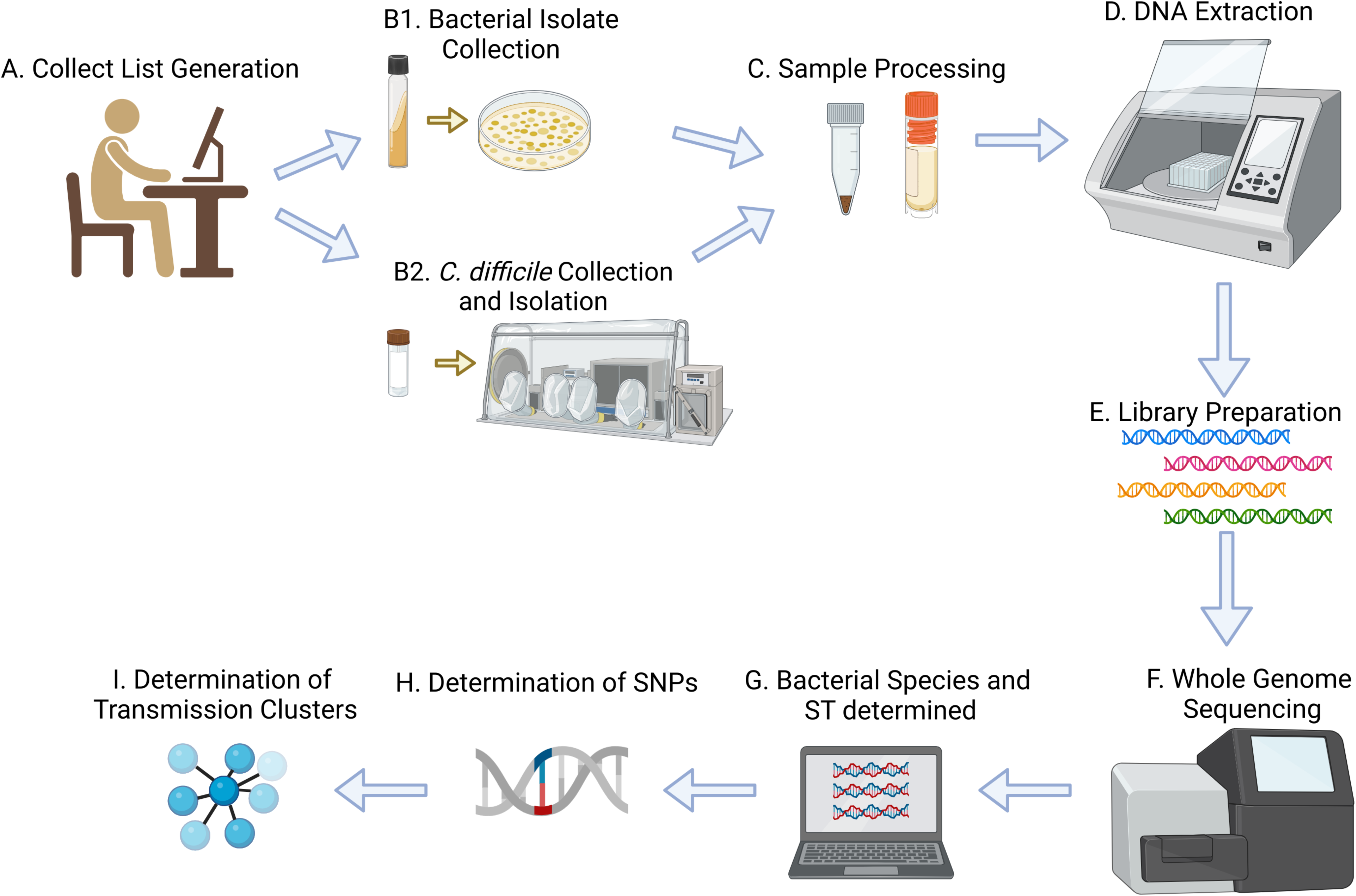
EDS-HAT real-time genomic surveillance methods. A) Biweekly collect list generation for all EDS-HAT organisms of interest obtained from patients admitted to the hospital for ≥ 3 days or had a previous hospital exposure in the prior 30 days, B1) Bacterial isolate collection from the UPMC Clinical Microbiology Laboratory, B2) Clinical stool specimen collection and *C. difficile* isolation using a Coy anaerobic chamber, C) Processing samples into pellets and glycerol stocks, D) DNA extraction, E) Library preparation using Eppendorf epMotion 5075, F) WGS using MiSeq or NextSeq550 (Phase 1); NextSeq1000 or NovaSeq X (Phase 2), G) Bioinformatic analysis to determine bacterial species and sequence type (ST), H) Determination of SNPs between isolates, and I) Determination of transmission clusters. The average turnaround time from MiGEL sample collection to determination of transmission clusters was 10 days. Created with BioRender.

We provide details about our methods for a three-year timeframe, after our initial optimization period, beginning in March 2022. We divided this timeframe into two distinct phases, represented by our transition from smaller instrumentation (MiSeq and NextSeq550) in Phase 1 before expanding our throughput and upgrading to the NextSeq1000 and NovaSeq X Plus in Phase 2.

#### Isolate collection

Bacterial samples were collected by MiGEL twice weekly at the CLB from pure cultures isolated from clinical specimens. (Fig 1.B1). CLB technologists subcultured all eligible gram-negative isolates from aerobic bacterial cultures to nutrient agar slants. We identified isolates of interest from the CLB, subcultured to trypticase soy agar with 5% sheep blood agar plates (BAP) (BD, Franklin Lakes, NJ), transported to MiGEL, and incubated at 37°C overnight in the presence of 5% CO_2_. The eligible gram-positive isolates from the CLB were transferred from one BAP to another and then transported and incubated at MiGEL following the same procedure. The following day, sample information was imported into the MiGEL database, and a de-identified specimen ID was generated per sample.

#### *Clostridioides difficile* collection and culture

We collected clinical stool specimens that tested positive for *C. difficile* by culture-independent diagnostic testing(18) and performed the following protocol to isolate this organism directly from the stool. In a biosafety cabinet, stool samples were subcultured onto cycloserine-cefoxitin-mannitol-agar with taurocholate and lysozyme (CCMA-TAL) plates to select for *C. difficile* growth. Plates were transferred into a Coy anaerobic chamber (Coy Laboratory Products, Grass Lake, MI) and incubated at 37°C for 48 hours. Colonies of *C. difficile* were passaged to a second CCMA plate and incubated at 37°C in the anaerobic chamber for an additional 24-48 hours. Isolates were confirmed as *C. difficile* by testing for L-Proline aminopeptidase production using a PRO Disc test (Remel, San Diego, CA; Fig 1B2).

### Sample preparation and DNA extraction

To begin sample preparation for WGS, microcentrifuge tubes containing 750 µL phosphate buffered saline (PBS) were inoculated with a quarter-portion of a 10 µL loop of bacteria (a half-portion was used for *C. difficile*) from the BAP or CCMA plate. The tubes were centrifuged at 6.0⨯*g* for 10 minutes to generate a pellet; the supernatant was removed using a P1000 pipette (Fig 1C). For samples not proceeding immediately to extraction, the pellets were stored at –20°C. Isolate stocks for long-term storage for all bacterial isolates (including *C. difficile*) were prepared by inoculating a 10 µL loop of bacteria into cryovials containing 1 mL of nutrient broth with 20% glycerol and then stored at –80°C.

The bacterial pellets were re-suspended in 500 µL PBS prior to extraction. DNA was extracted using the MagMAX DNA Multi-Sample Ultra 2.0 extraction kit on the KingFisher Apex (Thermo Fisher Scientific, Waltham, MA) per manufacturer’s instructions (Fig 1D). Briefly, this procedure isolates and purifies nucleic acids using magnetic bead-based technology. DNA was eluted in 100 µL of elution buffer and then quantified using a Qubit broad range dsDNA kit (Life Technologies, Carlsbad, CA). Samples with a concentration ≥3.5ng/µL were considered for WGS. For samples that did not meet this criterion, DNA was re-extracted.

### Library preparation

DNA libraries were prepared on an epMotion 5075t (Eppendorf, Hamburg, Germany) using a DNA Prep-(M)-Tagmentation kit (Illumina, San Diego, CA), utilizing half-volume reactions for BLT/TB1 and EPM reagents (Fig 1E). A unique 10-mer index adapter sequence was ligated to each sample (IDT, Coralville, IA). Briefly, this protocol uses bead-linked transposomes to tagment and amplify the adapter-tagged DNA segments. Eight individual libraries were pooled together by combining 5 µL into a single tube. Pooled libraries were quantified using a Qubit high sensitivity dsDNA kit per manufacturer’s protocol. The library pool was normalized to 4 nM with resuspension buffer (RSB), assuming a fragment size of 700 nucleotides. Additional pools were combined using equimolar concentration into a single tube. The distribution of the fragments in the final sequencing pool was assessed using an Agilent Tapestation D1000 or D5000 screen tape and reagents per manufacturer’s protocol (Agilent Technologies, Santa Clara, CA).

### Whole genome sequencing

#### Phase 1 (March 2022-July 2024)

DNA libraries were sequenced weekly using an Illumina MiSeq (≤32 samples on a v3, 600-cycle kit) or NextSeq550 (33-96 samples on a v2.5, 300-cycle kit) platform (Fig 1F). The pooled DNA library was denatured using 0.2N NaOH and spiked with 1% PhiX. The DNA library was diluted, using the average library length, to the final loading concentration of 16pM for the MiSeq or 1.5-1.6pM for the NextSeq550. A commercial lab was used for sequencing in rare instances where personnel were unavailable for in-house sequencing. For these occasions, DNA was extracted and sent for same-day delivery using a local medical courier service, followed by library preparation and sequencing at the commercial lab.

#### Phase 2 (July 2024-February 2025)

DNA libraries were sequenced using an Illumina NextSeq1000 (48-72 samples on a P1, XLEAP-SBS 300-cycle kit; 2⨯150 bp reads) or NovaSeq X Plus (73-192 samples on a v1, 300-cycle kit; 2⨯150 bp reads) platform (Fig 1F); the DNA library was spiked with 4% PhiX and diluted, using the average library length, to the final loading concentration of 850pM for the NextSeq1000 or 300pM for the NovaSeq X Plus. DNA libraries underwent onboard denaturation.

### Bioinformatics and data analysis

#### Sequencing data quality control (QC)

We developed a publicly available Python-based bioinformatics pipeline (https://github.com/mpgriffith/edshat-pipeline) that was executed weekly for newly sequenced samples. Before execution of the real-time bioinformatics pipeline, the data was first downloaded from BaseSpace Sequence Hub v7.18.0 (Illumina), sample reads were demultiplexed using bcl2fastq c2.20 (Illumina) software, and results were parsed into individual directories (S1 Fig). Specific to to the NovaSeq X Plus, image analysis and base calling were conducted by NovaSeq Control Software (NCS; Illumina).

#### Genome assembly and characterization

WGS reads were assembled using Unicycler v0.5.0 and annotated using Prokka v1.14(19). Multilocus sequence types (STs) were assigned using PubMLST typing schemes for all organisms except *Serratia* spp. and *Providencia* spp., which do not have ST schemes (mlst v2.11)(20). Reads were mapped using Kraken2 with the Kraken standard database to determine the most prevalent species(21).

#### Sequencing data quality control (QC)

Quast was used to determine the following QC attributes for each sample: total genome length, contig counts, and N50(22). If any sample contained a mixture of species identified in the read data, we attempted to rescue the sample by excluding reads attributed to species not indicated. For this, we used the “extract_kraken_reads.py” script from the KrakenTools package(23) to remove the reads mapping to the wrong genus. The remaining reads files were then re-analyzed with our pipeline. Isolates passed QC if 1) the most prevalent species by Kraken2 was the expected organism, 2) the assembly length was within 20% of the expected genome length, 3) the assembly was ≤ 350 contigs, and 4) there was at least 35⨯ depth (Fig 1G). Data were stored on our in-house server (Dell PowerEdge T640 with 172TB hard drive space, 1.48TB RAM and two Intel® Xeon® Gold 6240R Processor CPUs).

#### Determining genetically-related clusters and downstream applications

As part of the bioinformatics pipeline, samples from the most recent sequencing batch were compared against all previously collected isolates of the same species using single nucleotide polymorphisms (SNPs) to identify instances of genetically-related clusters. Pairwise SNPs were determined one of two methods (Fig 1H). i) Pairwise core genome SNPs (cgSNPs) were determined using Snippy v4.3.0, a reference-based method, for isolates with the same ST(24). SNP distances were calculated from the core alignment using ‘snp-dists’(25). ii) SKA v1.0, a reference-free method, was used to calculate SNP distances for isolates of the same species(26). We selected the minimum SNP distance for each pairwise comparison quantified by Snippy or SKA to determine clusters of genetically similar isolates. These clusters were defined using hierarchical clustering with average linkage and a cutoff of ≤15 or ≤10 SNPs for all species (the SNP threshold to infer transmission was modified in May 2024(11)) except *C. difficile*, for which a cutoff of ≤2 SNPs was used (Fig 1H; https://scipy.org/(10)). Phylogenetic trees were generated upon request using randomized accelerated maximum likelihood (RAxML; version 8.2.12)(27) using the general time reversible model of evolution (GTRCAT), Lewis correction for ascertainment bias, and 100 bootstrap replicates. We used AMRfinder to determine the presence of antimicrobial resistance gene sequences(28). The electronic health records (EHR) for patients with genetically similar isolates were reviewed to determine potential epidemiological links (sequencing data and transmission information was not deposited in patient EHR), and was reported to the IP&C team by email, who implemented targeted mitigation measures whenever possible for clusters with identified transmission routes.

### Cost analysis

A cost estimate for our optimized EDS-HAT real-time genomic surveillance methods was determined, standardized to 2025 US dollars, and included the cost of personnel, reagents, and supplies, and was analyzed comparatively for each sequencing platform used in this study (S1 Table). Equipment costs were not considered for weekly run costs and laboratories are assumed to have a basic setup, including typical molecular biology equipment and machines. Personnel costs (salary, including fringe benefits) were determined using the average pay scale of Laboratory Technician III (90% effort) and Bioinformatics Research Analyst II (50% effort) positions at the University of Pittsburgh, Pittsburgh, PA. Reagent and supply costs were determined using manufacturer pricing as of August 5, 2025.

## Results

### Weekly sequencing runs

From March 2022 to March 2025, MiGEL conducted real-time EDS-HAT, collecting and sequencing 7,813 bacterial isolates, with a weekly average sample count of *N*=60 or 80 (Phases 1 or 2, respectively) and an average genome size of 4.87 million nucleotides. The mean overall observed WGS failure rate was 4.5% (standard deviation, SD=0.005). The most common reasons for failure included low genome coverage (1.4%), mixed species or an unexpected species designation as determined by the clinical lab (0.9%), or poor assembly (2.2%). The most commonly sequenced organism was *Pseudomonas aeruginosa* (*N*=2,106) and the least sequenced was *Burkholderia* spp. (*N*=36; Table 1). The average turnaround time to complete the EDS-HAT workflow from MiGEL sample collection to bioinformatic analysis was approximately 10 days, with an average instrument run time of ∼25 and ∼16 hours for NextSeq550 and NextSeq1000, respectively; S2 Table).

**Table 1.** EDS-HAT organisms sequenced by MiGEL (March 1, 2022 to February 28, 2025).

| Organism | Genome Size (Mb) | Count Sequenced by MiGEL | Failures |  |  |
| --- | --- | --- | --- | --- | --- |
|  |  |  | Count | % by Species | % of Total |
| *Acinetobacter* species | 3.9 | 172 | 8 | 4.65 | 0.10 |
| *Burkholderia* species | 7.2 | 36 | 6 | 16.67 | 0.08 |
| *Clostridioides difficile* | 4.2 | 468 | 34 | 7.26 | 0.44 |
| *Citrobacter* species | 4.7 | 156 | 5 | 3.21 | 0.06 |
| *Enterobacter cloacae* | 5.3 | 138 | 2 | 1.45 | 0.03 |
| *Escherichia coli* | 5.3 | 433 | 8 | 1.85 | 0.10 |
| *Klebsiella oxytoca* | 5.9 | 74 | 0 | 0 | 0 |
| *Klebsiella pneumoniae* | 5.3 | 553 | 7 | 1.27 | 0.09 |
| MRSA | 2.9 | 1,077 | 24 | 2.23 | 0.31 |
| *Proteus* species | 4.06 | 934 | 17 | 1.82 | 0.22 |
| *Providencia* species | 4.5 | 103 | 2 | 1.94 | 0.03 |
| *Pseudomonas aeruginosa* | 6.7 | 2,106 | 42 | 1.99 | 0.54 |
| *Pseudomonas* species | 6.6 | 86 | 9 | 10.47 | 0.12 |
| *Serratia* species | 5.2 | 532 | 13 | 2.44 | 0.17 |
| *Stenotrophomonas* species | 4.8 | 356 | 20 | 5.62 | 0.26 |
| VRE | 2.9 | 589 | 7 | 1.19 | 0.09 |
| **GRAND TOTAL** |  | **7,813** | **204** | **-** | **2.61** |

Bacterial genome sizes and the grand total of each organism collected for sequencing is tabulated. Isolates failing genome quality control are tabulated. Mb, megabase.

### Sample count

To estimate the maximum number of samples per sequencing flow cell, we considered our weekly average genome size and approximated a minimum target of 80× coverage per sample. For Phase 1, we considered a maximum of 32 or 96 samples for MiSeq or NextSeq550, respectively, and for Phase 2, a maximum of 72 or 192 samples for NextSeq1000 or NovaSeq X Plus, respectively. These reported maximum sample counts are slightly lower than the actual maximum counts to allow room for genome variability in the sequencing pool. When sequencing pools that contained a greater number of organisms with a smaller average genome size, we were able to increase the number of isolates per flow cell without compromising run quality or per organism genome coverage (Fig 2). Based on the MiGEL average genome size, and to maximize cost savings, runs containing 32-40 samples were sequenced on the MiSeq, and runs containing 40-96 samples were sequenced on the NextSeq550 platform during Phase 1. For Phase 2, runs containing 48-72 samples were sequenced on the NextSeq1000 and runs containing 72-192 samples were sequenced on the NovaSeq X Plus. During this study, 18 runs were performed on the MiSeq, 90 on the NextSeq550, 9 on the NextSeq1000, and 17 on the NovaSeq X Plus platforms.

**Fig 2.**
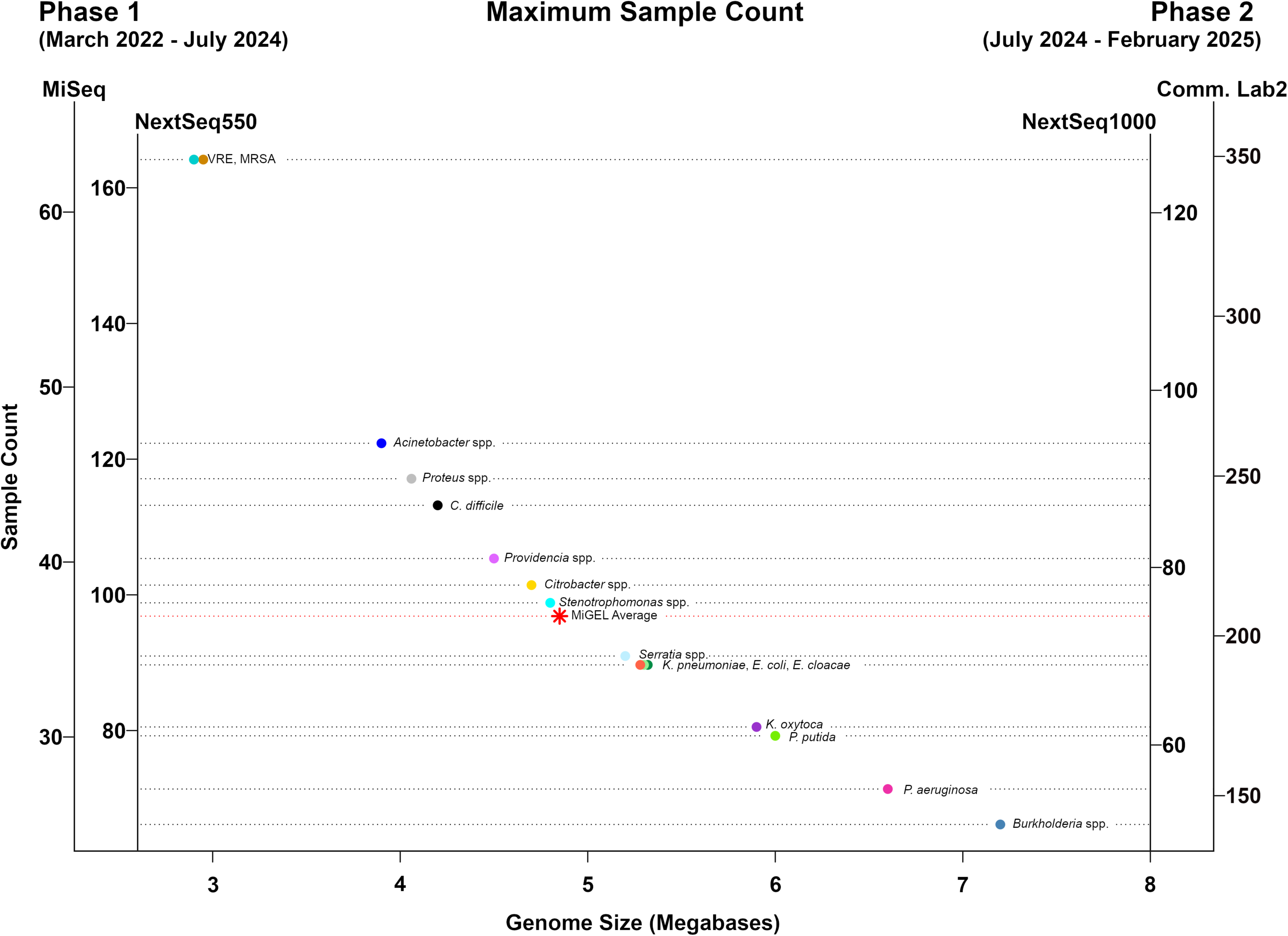
Maximum number of genomes that can be sequenced on the MiSeq v3 600 cycle flow cell or NextSeq550 mid-output v2.5 300 cycle flow cell (Phase 1) compared to NextSeq1000 P1 XLEAP-SBS 300 cycle flow cell or NovaSeq X flow cell (Phase 2) based on size (Mb). Sample counts were calculated using Illumina coverage calculator based on 80x coverage criteria. Genome size of an average run by MiGEL is shown in comparison to individual organism sizes (red star).

### Data output

For an average Phase 1 run of 60 samples on the NextSeq550, MiGEL observed a maximum output of 52 Gb of data and an average of 100 million reads (S2 Table). For an average Phase 2 run of 80 samples on the NovaSeq X Plus platform, MiGEL observed a maximum output of 196 Gb data and an average of 402 million reads.

### Bioinformatic analyses run time

We performed bioinformatics analyses using 40 CPU cores; the time required to completion was approximately 44 h (*N*=72 samples). This time estimate included a per sample comparison to other samples of the same species in our EDS-HAT database (Table 1). The times required for each step or program are detailed in Supplemental Table 3.

### Cost analysis

The cost to run real-time EDS-HAT weekly was categorized into sample processing, DNA extraction and quantification, library preparation, flow cell cost, and personnel (Table 2). The programs used for the bioinformatic analyses were freely downloadable, therefore our only upfront cost was an investment in a server.

**Table 2.** Weekly cost estimates per sample (March 2022–February 2025).

|  | Phase 1: March 2022 – July 2024 |  |  |  |  | Phase 2: July 2024 – February 2025 |  |  |  |  |
| --- | --- | --- | --- | --- | --- | --- | --- | --- | --- | --- |
| Method | MiSeq Cost | <u>NextSeq550 Cost</u> |  |  | Commercial Lab 1 Sequencing Cost<sup>#</sup> | NextSeq 1000 Cost | <u>Commercial Lab 2 (NovaSeq X Plus) Cost</u> |  |  | Commercial Lab 1 Sequencing Cost<sup>#</sup> |
|  | 32 samples | 48 samples | 60* samples | 96 samples | ≤32 samples | 72 samples | 80* samples | 96 samples | 192 samples | ≤32 samples |
| Sample Processing | \$2.25 | \$2.25 | \$2.25 | \$2.25 | \$2.25 | \$2.25 | \$2.25 | \$2.25 | \$2.25 | \$2.25 |
| DNA Extraction & Quantification | \$9.35 | \$8.66 | \$8.39 | \$7.98 | \$9.35 | \$8.21 | \$8.11 | \$7.98 | \$7.98 | \$9.35 |
| Library Preparation | \$22.54 | \$21.78 | \$21.47 | \$21.01 | \$75 | \$21.26 | \$21.16 | \$21.01 | \$21.01 | \$65 |
| Sequencing Kit (Flow Cell) | \$58.47 | \$43.13 | \$34.50 | \$21.56 | | \$13.89 | \$14.65 | \$12.21 | \$6.10 | |
| LABORATORY COST PER SAMPLE | \$92.61 | \$75.82 | \$66.61 | \$52.80 | \$86.60 | \$45.61 | \$46.17 | \$43.45 | \$37.34 | \$76.60 |
| Personnel |  |  |  |  |  |  |  |  |  |  |
| -laboratory technician, 90% effort (\$970/week) | \$30.31 | \$20.21 | \$16.17 | \$10.10 | \$30.31 | \$13.47 | \$12.13 | \$10.10 | \$5.05 | \$30.31 |
| -bioinformatician, 50% effort (\$524/week) | \$16.38 | \$10.92 | \$8.73 | \$5.46 | \$16.38 | \$7.28 | \$6.55 | \$5.46 | \$2.73 | \$16.38 |
| TOTAL COST PER SAMPLE | \$139.30 | \$106.95 | \$91.51 | \$68.36 | \$133.29 | \$66.36 | \$64.85 | \$59.01 | \$45.12 | \$123.29 |

<sup>#</sup>used for comparison only, not included in calculations for averages;
\*average sample count per phase.

#### Phase 1

The lowest laboratory cost per sample ($53) was achieved when the maximum number of samples (*N*=96) was sequenced using the NextSeq550 platform. Costs ranged from $53-$93 per sample, depending on the platform and sample counts.

#### Phase 2

The lowest laboratory cost per sample ($40) was achieved when 192 samples were sequenced using the NovaSeqX Plus platform. Costs ranged from $37-$46 per sample.

We determined a sample count cutoff, per phase, to decide which sequencer to use for each run. For Phases 1 or 2, the sample count cutoffs were 40 or 72 samples, respectively (Fig 3). There was an inverse relationship, in both phases, between the number of samples sequenced and flow cell cost, as per sample costs significantly decreased when a greater number of samples were multiplexed on the appropriate flow cell.

**Fig 3.**
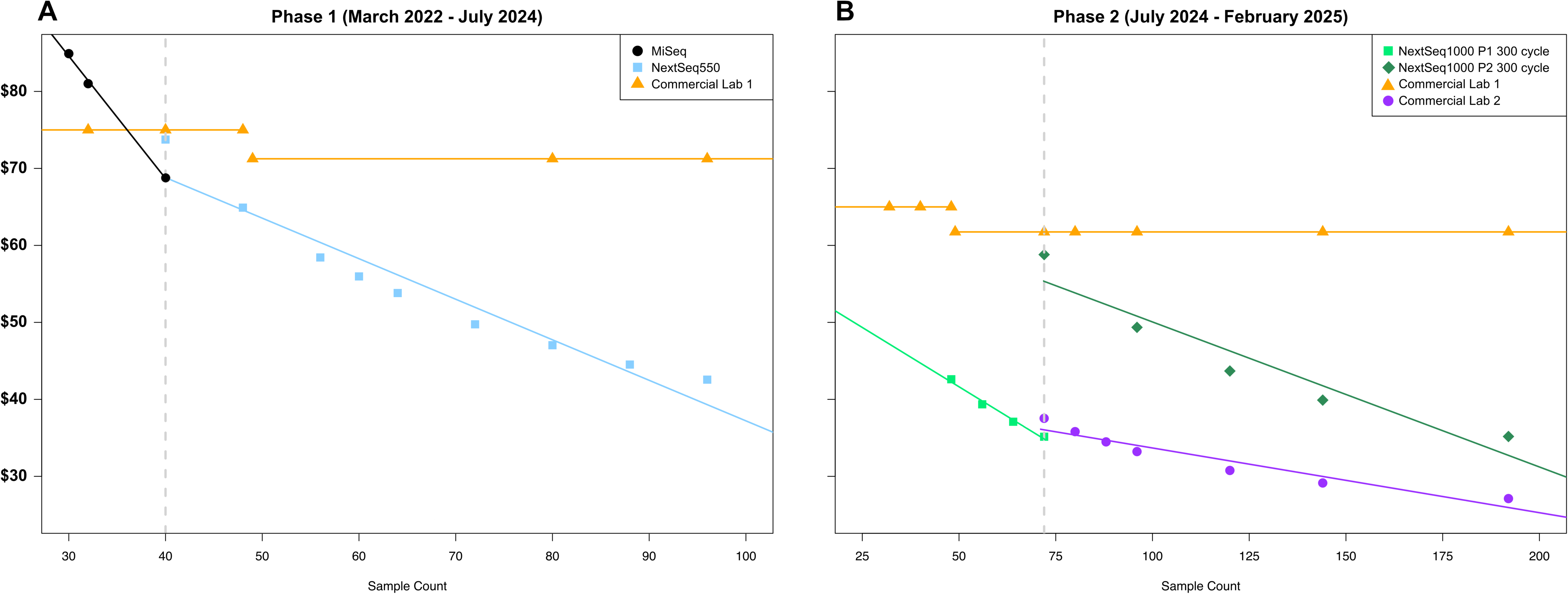
Whole genome sequencing cost per sample comparisons. A) MiSeq, NextSeq550, and commercial laboratory 1. Throughput cutoff between platforms is shown at *N*=40 samples (gray dashed line). B) NextSeq1000 P1 300 cycle and P2 300 cycle flow cells, Commercial laboratory 1 and Commercial laboratory 2. Throughput cutoff between platforms is shown as *N*=72 samples (gray dashed line). Commercial lab 1 used by MiGEL offers a discount of 5% for orders ≥ 48 samples (as of September 2025). Data points represent instances of cost by sample count and lines of best fit are shown for each sequencing method.

The estimated weekly cost of personnel, based on the pre-tax salaries for one lab technician and one bioinformatician considering percent efforts and fringe benefits, totaled $1,494 ($970 and $524, respectively). When these costs were tabulated, the Phase 1 cost to run EDS-HAT on an average week of sequencing 60 samples totaled $5,491 ($285,511 per year), while Phase 2 costs totaled $5,188 for an average week of sequencing 80 samples ($269,755 per year).

## Discussion

In this study, we detailed an efficient laboratory workflow, our approach for bioinformatics analyses, and estimated the cost associated with implementing real-time WGS surveillance for pathogenic bacteria in a hospital system. EDS-HAT began in 2016 as a retrospective study(10) and, once we demonstrated the superiority of the system over traditional approaches, transitioned in November 2021 to a real-time workflow, subsequent bioinformatic analyses, and reporting of results to the hospital IP&C team(11). MiGEL has been performing prospective WGS surveillance for multiple organisms in real time for the UPMC hospital system for over four years. By doing so, our hospital system has dramatically changed the way outbreaks are being detected and interrupted (10, 11, 29).

We provide details about our methods for a three-year timeframe, after our initial optimization period, beginning in March 2022. We divided this timeframe into two distinct phases, represented by our transition from smaller instrumentation (MiSeq and NextSeq550) in Phase 1 before expanding our throughput and upgrading to the NextSeq1000 and NovaSeq X Plus in Phase 2. We determined that the per sample cost for WGS ranged from $53 to $93 (average week: $67; Phase 1), and $37 to $46 (average week: $46; Phase 2). Furthermore, with the addition of staff salaries, the weekly cost for an average week of real-time sequencing and analyses was $5,491 (*N*=60 samples; Phase 1) that decreased to $5,188 (*N*=80 samples; Phase 2). Our lower cost was achieved, in part, by increasing sample counts per flow cell while utilizing the appropriate instrument and by using half-volumes of some reagents during library preparation. The transition to larger sequencing instruments also provided the use of more efficient sequencing chemistry technologies.

During Phase 1, we showed an estimated cost savings of $79,227–$285,160 per year by implementing a real-time WGS surveillance system, which was based on an average cost of $71 for sample preparation and sequencing (adjusted to 2025 USD)(29). Since then, we have made further improvements in Phase 2 (average costs were $48/sample on the NextSeq1000 [SD, $2.28], and $45/sample on the NovaSeq X Plus [SD, $3.50]). 10/17/2025 11:58:00 AM

A major concern of hospitals with implementing programs like EDS-HAT is high cost. While sequencing was expensive years ago, the cost has significantly decreased over time(30). Furthermore, the cost of treating preventable hospital infections is high, and, in fact, EDS-HAT has been shown to be cost saving(29, 31). In addition, the associated laboratory and bioinformatics methods have become more streamlined, automatable, and efficient. With our quick turnaround time from the day the sample is collected by MiGEL, we provide evidence of outbreaks that helps to guide our hospital IP&C team to implement interventions. Taken together, this approach can identify important, otherwise-undetected outbreaks in a cost-effective manner—suggesting that WGS surveillance should eventually become standard practice in hospitals. More importantly, stopping transmission events quickly at the first sign of an outbreak cluster has the potential to reduce further spread of the infection and thus, reduce patient morbidity and mortality.

To accompany our methods, we computed Phase 2 cost per sample, including staff salaries, as $65 on an average week (range, $45-$66). This cost is specific for our hospital and could be different at other locations as summarized by Price and colleagues, who determined that the cost to perform WGS varies by country and city(32). For example, the prior cost-per-sample study, when converted to 2025 USD, showed the per sample sequencing cost ranged from approximately $59-$387 for the US and Italy, respectively. In this study, we determined our current average per sample cost (without considering staff salaries, for comparison) was $44. The primary factors in determining this cost estimate were sample counts per run and average organism genome size. For reference, we provide the maximum number of samples that can be sequenced on either MiSeq, NextSeq550, NextSeq1000, or NovaSeq X Plus platforms by organism, considering genome size, along with the average genome size sequenced over one year by MiGEL (Fig 2). In addition, Fig 3 shows that a sample count of 40 or 72 are appropriate cutoffs to decide which sequencing platform to use for Phase 1 and Phase 2 platforms, respectively, while maintaining sufficient genome coverage. Generally, we find sequencing more samples at once reduced the cost of sequencing per sample, with the exception of utilizing specific commercial labs. While one commercial lab offered a discounted price once the sample count reached 48, we found the fixed price was overall more costly than performing in-house sequencing (Fig 3). Furthermore, we found sequencing costs decreased over time. MiGEL estimated a $72 average per sample cost in 2021, which dropped to $65 in 2023 and is now further reduced to $37 in 2025.

We note limitations with this study. First, the reagent and supply costs presented in this manuscript represent discounted pricing provided to our university from some manufacturers. Other institutions could be charged differently, which will alter the costs. Second, MiGEL uses robotic instruments for nucleic acid extractions and library preparation, which helps save time and decrease pipetting errors on the bench. Some institutions may not have such instruments and will need to accommodate the laboratory methods accordingly; however, we do not think this represents a significant detriment to the process. Third, we only considered Illumina-based technology for this study. Other short-read sequencing technologies or long-read sequencing were not assessed. Fourth, we have demonstrated how we made our method cost-effective for us at an academic, tertiary hospital. These estimates may not translate to smaller hospitals. Moreover, EDS-HAT would run optimally if WGS was performed on all clinical isolates, including colonization(33); however, specific species and clinical cultures were prioritized due to budgetary limitations. Finally, we achieved our lowest cost outsourcing to a NovaSeq X Plus sequencer, housed at a commercial laboratory. Budgetary considerations restricted our ability to purchase this machine; thus, the instrument’s run time and select WGS metrics are unknown and therefore not included in this analysis.

In conclusion, we have shown the methods and our costs to implement a real-time WGS surveillance program. Healthcare institutions wishing to do the same could potentially identify outbreaks that would otherwise be missed. Further adoption of this approach has the potential to significantly enhance patient safety.

## Ethical approval

The University of Pittsburgh institutional review board provided ethics approval for this study.

## Data Availability

All data produced in the present study are available upon reasonable request to the authors.

https://www.ncbi.nlm.nih.gov/bioproject

https://github.com/mpgriffith/edshat-pipeline

## Acknowledgements

The authors would like to thank SeqCenter and Azenta for their assistance with sequencing. We thank the leaders and staff of the UPMC Clinical Laboratories, especially Tung Phan, MD, PhD, D(ABMM), Hannah Creager PhD, D(ABMM), and all members of the UPMC Presbyterian/Shadyside Infection Prevention & Control Team, especially Graham Snyder, MD and Ashley Ayres, MBA, CIC for their continued support. We also thank Jane Marsh, PhD, for her contributions to the EDS-HAT project. This publication made use of the PubMLST website (https://pubmlst.org/) developed by Keith Jolley (Jolley & Maiden 2010, BMC Bioinformatics, 11:595) and sited at the University of Oxford. The development of that website was funded by the Wellcome Trust.

## Supporting information

**S1 Table.** Vendor information and the reagents used to perform whole genome sequencing of bacterial isolates.

**S2 Table. Phase 1 (2a)**. Sequencing metrics per platform using the average number of samples (*N*=60) sequenced by MiGEL. **Phase 2 (2b)**. Sequencing metrics per platform using the average number of samples (*N*=80) sequenced by MiGEL.

**S3 Table.** Bioinformatics pipeline performance metrics.

**S1 Figure**. EDS-HAT bioinformatics pipeline.

## References

1. Finance AA. Inflation Calculator. 2025 [Available from: https://www.officialdata.org/us/inflation/.

2. RD. S. The Direct Medical Costs of Healthcare-Associated Infections in U.S. Hospitals and the Benefits of Prevention. In: Prevention CfDCa, editor. 2009.

3. Mustapha MM, Srinivasa VR, Griffith MP, Cho ST, Evans DR, Waggle K, et al. Genomic Diversity of Hospital-Acquired Infections Revealed through Prospective Whole-Genome Sequencing-Based Surveillance. mSystems. 2022;7(3):e0138421.

4. Sherry NL, Gorrie CL, Kwong JC, Higgs C, Stuart RL, Marshall C, et al. Multi-site implementation of whole genome sequencing for hospital infection control: A prospective genomic epidemiological analysis. Lancet Reg Health West Pac. 2022;23:100446.

5. Ward DV, Hoss AG, Kolde R, van Aggelen HC, Loving J, Smith SA, et al. Integration of genomic and clinical data augments surveillance of healthcare-acquired infections. Infect Control Hosp Epidemiol. 2019;40(6):649–55.

6. Neoh HM, Tan XE, Sapri HF, Tan TL. Pulsed-field gel electrophoresis (PFGE): A review of the “gold standard” for bacteria typing and current alternatives. Infect Genet Evol. 2019;74:103935.

7. Quainoo S, Coolen JPM, van Hijum S, Huynen MA, Melchers WJG, van Schaik W, et al. Whole-Genome Sequencing of Bacterial Pathogens: the Future of Nosocomial Outbreak Analysis. Clin Microbiol Rev. 2017;30(4):1015–63.

8. Mellmann A, Bletz S, Boking T, Kipp F, Becker K, Schultes A, et al. Real-Time Genome Sequencing of Resistant Bacteria Provides Precision Infection Control in an Institutional Setting. J Clin Microbiol. 2016;54(12):2874–81.

9. Sundermann AJ, Babiker A, Marsh JW, Shutt KA, Mustapha MM, Pasculle AW, et al. Outbreak of Vancomycin-resistant Enterococcus faecium in Interventional Radiology: Detection Through Whole-genome Sequencing-based Surveillance. Clin Infect Dis. 2020;70(11):2336–43.

10. Sundermann AJ, Chen J, Kumar P, Ayres AM, Cho ST, Ezeonwuka C, et al. Whole-Genome Sequencing Surveillance and Machine Learning of the Electronic Health Record for Enhanced Healthcare Outbreak Detection. Clin Infect Dis. 2022;75(3):476–82.

11. Sundermann AJ, Kumar P, Griffith MP, Waggle KD, Rangachar Srinivasa V, Raabe N, et al. Real-Time Genomic Surveillance for Enhanced Healthcare Outbreak Detection and Control: Clinical and Economic Impact. Clin Infect Dis. 2025.

12. Didelot X, Bowden R, Wilson DJ, Peto TEA, Crook DW. Transforming clinical microbiology with bacterial genome sequencing. Nature reviews Genetics. 2012;13(9):601–12.

13. Neumann B, Bender JK, Maier BF, Wittig A, Fuchs S, Brockmann D, et al. Comprehensive integrated NGS-based surveillance and contact-network modeling unravels transmission dynamics of vancomycin-resistant enterococci in a high-risk population within a tertiary care hospital. PLoS One. 2020;15(6):e0235160.

14. Raabe NJ, Valek AL, Griffith MP, Mills E, Waggle K, Srinivasa VR, et al. Real-time genomic epidemiologic investigation of a multispecies plasmid-associated hospital outbreak of NDM-5-producing Enterobacterales infections. Int J Infect Dis. 2024;142:106971.

15. Sundermann AJ, Rangachar Srinivasa V, Mills EG, Griffith MP, Waggle KD, Ayres AM, et al. Two Artificial Tears Outbreak-Associated Cases of Extensively Drug-Resistant Pseudomonas aeruginosa Detected Through Whole Genome Sequencing-Based Surveillance. J Infect Dis. 2024;229(2):517–21.

16. Sundermann AJ, Griffith M, Rangachar Srinivasa V, Ereifej D, Waggle K, Van Tyne D, et al. Environmental contamination of postmortem blood cultures detected by whole-genome sequencing surveillance. Infect Control Hosp Epidemiol. 2023;44(12):2103–5.

17. Sundermann AJ, Chen J, Miller JK, Saul MI, Shutt KA, Griffith MP, et al. Outbreak of Pseudomonas aeruginosa Infections from a Contaminated Gastroscope Detected by Whole Genome Sequencing Surveillance. Clin Infect Dis. 2021;73(3):e638–e42.

18. Crobach MJ, Planche T, Eckert C, Barbut F, Terveer EM, Dekkers OM, et al. European Society of Clinical Microbiology and Infectious Diseases: update of the diagnostic guidance document for Clostridium difficile infection. Clin Microbiol Infect. 2016;22 Suppl 4:S63–81.

19. Seemann T. Prokka: rapid prokaryotic genome annotation. Bioinformatics. 2014;30(14):2068–9.

20. Seemann T. MLST. https://github.com/tseemann/mlst. 2025.

21. Wood DE, Lu J, Langmead B. Improved metagenomic analysis with Kraken 2. Genome Biol. 2019;20(1):257.

22. Gurevich A, Saveliev V, Vyahhi N, Tesler G. QUAST: quality assessment tool for genome assemblies. Bioinformatics. 2013;29(8):1072–5.

23. Lu J, Rincon N, Wood DE, Breitwieser FP, Pockrandt C, Langmead B, et al. Metagenome analysis using the Kraken software suite. Nat Protoc. 2022;17(12):2815–39.

24. Seemann T. Snippy. https://github.com/tseemann/snippy. 2025.

25. Seemann T. SNP-dists. https://github.com/tseemann/snp-dists. 2025.

26. SR. H. SKA: Split Kmer Analysis Toolkit for Bacterial Genomic Epidemiology. . 2018.

27. Stamatakis A. RAxML version 8: a tool for phylogenetic analysis and post-analysis of large phylogenies. Bioinformatics. 2014;30(9):1312–3.

28. Feldgarden M, Brover V, Gonzalez-Escalona N, Frye JG, Haendiges J, Haft DH, et al. AMRFinderPlus and the Reference Gene Catalog facilitate examination of the genomic links among antimicrobial resistance, stress response, and virulence. Sci Rep. 2021;11(1):12728.

29. Kumar P, Sundermann AJ, Martin EM, Snyder GM, Marsh JW, Harrison LH, et al. Method for Economic Evaluation of Bacterial Whole Genome Sequencing Surveillance Compared to Standard of Care in Detecting Hospital Outbreaks. Clin Infect Dis. 2021;73(1):e9–e18.

30. Wetterstrand K. DNA Sequencing Costs: Data from the NHGRI Genome Sequencing Program (GSP). 2025 [Available from: www.genome.gov/sequencingcostsdata.

31. Gordon LG, Elliott TM, Forde B, Mitchell B, Russo PL, Paterson DL, et al. Budget impact analysis of routinely using whole-genomic sequencing of six multidrug-resistant bacterial pathogens in Queensland, Australia. BMJ Open. 2021;11(2):e041968.

32. Price V, Ngwira LG, Lewis JM, Baker KS, Peacock SJ, Jauneikaite E, et al. A systematic review of economic evaluations of whole-genome sequencing for the surveillance of bacterial pathogens. Microb Genom. 2023;9(2).

33. Sundermann AJ, Rangachar Srinivasa V, Mills EG, Griffith MP, Evans E, Chen J, et al. Genomic sequencing surveillance of patients colonized with vancomycin-resistant Enterococcus (VRE) improves detection of hospital-associated transmission. medRxiv. 2024.

