## Supplementary material for "Methods for cost-efficient, whole genome sequencing surveillance for enhanced detection of outbreaks in a hospital setting": S1 Table

**Supplementary Table 1.**

Vendor information and the reagents used to perform whole genome sequencing of bacterial isolates.

| Vendor | Item Number | Item Description |
| --- | --- | --- |
| Thermo Fisher | A36570 | MagMax DNA Multi-Sample Ultra 2.0 DNA Extraction Kit |
| Thermo Fisher | Q32853 | Qubit broad range dsDNA Quantification kit |
| Illumina | 20060059 | DNA Prep Kit |
| Integrated DNA Technologies (IDT) | - | Nextera Custom Unique Dual Indexes |
| Illumina | 20091654 | Illumina DNA/RNA UD Indexes Set A, Tagmentation (96 indexes) |
| Illumina | MS-102-3003 | MiSeq Reagent Kit v3 (600 cycles) |
| Illumina | 20024905 | NextSeq 500/550 Mid Output Kit v2.5 (300 cycles) |
| Illumina | 20050264 | NextSeq1000/2000 P1 XLEAP-SBS Reagent Kit (300 cycles) |
| Illumina | FC-110-3001 | PhiX Control |
