## Supplementary material for "Methods for cost-efficient, whole genome sequencing surveillance for enhanced detection of outbreaks in a hospital setting": S2 Table

**Supplementary Table 2a.**

**Phase 1.** Sequencing metrics per platform using the average number of samples (*N*=60) sequenced by MiGEL.

|  | MiSeq^a^ | NextSeq550^b^ | MiGEL  (60 samples, NextSeq550) |
| --- | --- | --- | --- |
| Run Time | 4-56 hours | 12-30 hours | 25 hours |
| Maximum Read Output | 15 Gb | 120 Gb | 52 Gb |
| Average Read Output | 24 million | 260 million | 100 million |
| Maximum Read Length | 2 x 300 bp | 2 x 150 bp | 2 x 150 bp |

a.illumina.com

b.https://med.stanford.edu/sfgf/pricing.html

**Supplementary Table 2b.**

**Phase 2.** Sequencing metrics per platform using the average number of samples (*N*=80) sequenced by MiGEL.

|  | NextSeq1000 | NovaSeq X Plus | MiGEL  (80 samples, NovaSeq X) |
| --- | --- | --- | --- |
| Run Time | 16 – 19 hours | Not provided^#^ | 16 hours |
| Maximum Read Output | 43 Gb | 196 Gb | 124 Gb |
| Average Read Output | 112 million | 402 million | 143 million |
| Maximum Read Length | 2 x 150 bp | 2 x 150 bp | 2 x 150 bp |

^#^Expected turnaround time for NovaSeq X Plus (Commercial Lab 2) is 5 business days or less from sample receipt
