## Supplementary material for "Methods for cost-efficient, whole genome sequencing surveillance for enhanced detection of outbreaks in a hospital setting": S3 Table

**Supplementary Table 3. Bioinformatics pipeline performance metrics.** The time (minutes) and memory (RAM; MB, megabyte) usage statistics for the steps described for genomic surveillance. Of note, multithreaded steps (Unicycler, Prokka, QUAST, SKA compare scripts, AMRFinder, or RAxML) were run using 8 threads. SD, standard deviation.

Unicycler, bacterial genomes assembly tool.

Prokka (Prokaryotic genome annotation), annotates bacterial genomes.

PubMLST (Multi-Locus Sequence Typing), characterizes and classifies bacterial genomes using molecular typing.

Kraken2, taxonomic classification tool.

QUAST (QUality ASsessment Tool), evaluates quality of genome assemblies.

SKA FASTQ (all)/SKA (compare), determines single nucleotide polymorphisms (SNP) using Split K-mer Analysis (SKA), which compares bacterial genomes and determines genomic variants among species (reference-free variant calling). *SKA FASTQ (all)* generates a split K-mer file from the raw reads. *SKA* *(compare)* determines variants among specific isolates of interest using the split K-mer file.

Snippy (all)/ Snippy (core), compares bacterial genomes per Sequence Type (ST) and determines genomic variants (reference-based variant calling). *Snippy (all)* determines variants between one isolate and the reference genome (pairwise comparison). *Snippy (core)* determines variants that are common to all isolates, between ≥2 isolates and the reference genome.

Snippy (post), generates SNP matrices using the SNP alignment; for STs containing >20 genomes, the SNP matrix considers positions in ≥95% genomes that are contained within the core SNP alignment.

Combine SNPs, determines the minimum SNP distances per genome pairs by choosing either SKA or Snippy output.

Clusters, determines putative transmission clusters using hierarchical clustering with average linkage and a ≤10 SNP difference threshold.

Phylogenetic Tree, generated using Randomized Accelerated Maximum Likelihood (RAxML).

AMRFinder, identifies genomic elements related to bacterial resistance/virulence.

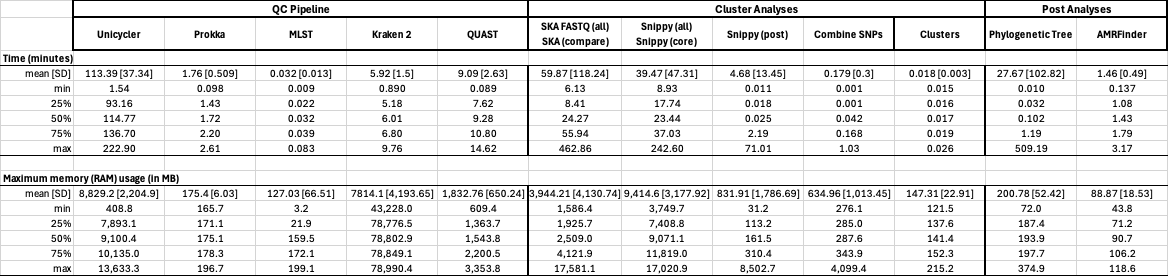
