## Supplementary figures and images for "Methods for cost-efficient, whole genome sequencing surveillance for enhanced detection of outbreaks in a hospital setting"

### S1 Fig

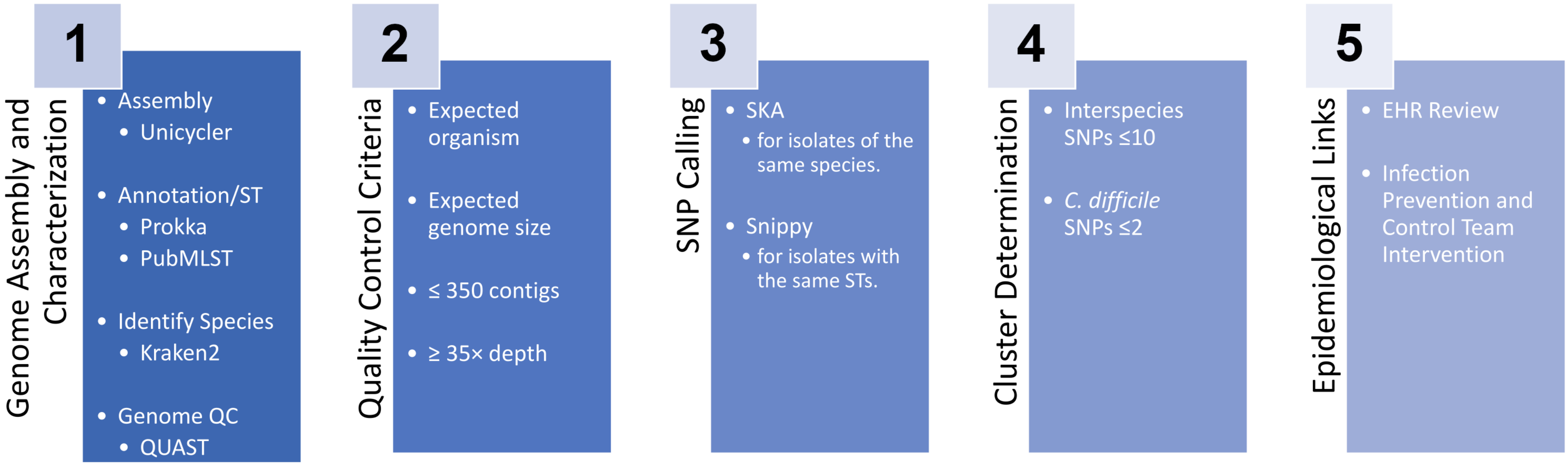
